# Mondo: Unifying diseases for the world, by the world

**DOI:** 10.1101/2022.04.13.22273750

**Authors:** Nicole A Vasilevsky, Nicolas A Matentzoglu, Sabrina Toro, Joseph E Flack, Harshad Hegde, Deepak R Unni, Gioconda F Alyea, Joanna S Amberger, Larry Babb, James P Balhoff, Taylor I Bingaman, Gully A Burns, Orion J Buske, Tiffany J Callahan, Leigh C Carmody, Paula Carrio Cordo, Lauren E Chan, George S Chang, Sean L Christiaens, Louise C Daugherty, Michel Dumontier, Laura E Failla, May J Flowers, H. Alpha Garrett, Jennifer L Goldstein, Dylan Gration, Tudor Groza, Marc Hanauer, Nomi L Harris, Jason A Hilton, Daniel S Himmelstein, Charles Tapley Hoyt, Megan S Kane, Sebastian Köhler, David Lagorce, Abbe Lai, Martin Larralde, Antonia Lock, Irene López Santiago, Donna R Maglott, Adriana J Malheiro, Birgit H M Meldal, Monica C Munoz-Torres, Tristan H Nelson, Frank W Nicholas, David Ochoa, Daniel P Olson, Tudor I Oprea, David Osumi-Sutherland, Helen Parkinson, Zoë May Pendlington, Ana Rath, Heidi L Rehm, Lyubov Remennik, Erin R Riggs, Paola Roncaglia, Justyne E Ross, Marion F Shadbolt, Kent A Shefchek, Morgan N Similuk, Nicholas Sioutos, Damian Smedley, Rachel Sparks, Ray Stefancsik, Ralf Stephan, Andrea L Storm, Doron Stupp, Gregory S Stupp, Jagadish Chandrabose Sundaramurthi, Imke Tammen, Darin Tay, Courtney L Thaxton, Eloise Valasek, Jordi Valls-Margarit, Alex H Wagner, Danielle Welter, Patricia L Whetzel, Lori L Whiteman, Valerie Wood, Colleen H Xu, Andreas Zankl, Xingmin Aaron Zhang, Christopher G Chute, Peter N Robinson, Christopher J Mungall, Ada Hamosh, Melissa A Haendel

## Abstract

There are thousands of distinct disease entities and concepts, each of which are known by different and sometimes contradictory names. The Monarch Initiative aims to integrate genotype, phenotype, and disease knowledge from a large variety of sources in support of improved diagnostics and mechanism discovery through various algorithms and tools. However, the lack of a unified system for managing disease entities poses a major challenge for both machines and humans to predict causes and treatments for disease. The multitude of disease resources have not been well coordinated nor computationally integrated. Furthermore, the classification of phenotypes and their association with diseases is another source of disagreement across sources. The Human Phenotype Ontology has helped to standardize phenotypic features across knowledge sources, but there was no equivalent computationally-harmonized disease ontology. To address these problems, a community of disease resources worked together to create the Mondo Disease Ontology as an open, community-driven ontology that integrates key medical and biomedical terminologies and is iteratively and regularly updated via manual curation and through synchronization with external sources using a Bayesian algorithm. Mondo supports disease data integration to improve diagnosis, treatment, and translational research. It records the sources of all data and is continually updated, making it suitable for research and clinical applications that require up-to-date disease knowledge.

**Research in Context:** *Evidence before this study:* Many disease terminologies currently exist, but there is not a definitive standard for encoding diseases while addressing requirements for information exchange. Existing sources of disease definitions include the National Cancer Institute Thesaurus (NCIt), the Online Mendelian Inheritance in Man (OMIM), Orphanet, SNOMED CT, Disease Ontology (DO), ICD-10, MedGen, and numerous others. Each of these is designed for a particular purpose, and as such has different strengths. However, these standards only partially overlap and often conflict in the classification or mapping approach, making it difficult to align them with each other and/or with other knowledge sources. This need to integrate information has resulted in a proliferation of mappings between disease entries in different resources; these mappings lack completeness, accuracy, and precision, and are often inconsistent between resources.

*Added value of this study:* In order to computationally leverage the available knowledge sources for diagnostics and to reveal underlying mechanisms of diseases, we need to understand which terms are meaningfully equivalent across different resources. This will allow integration of associated information, such as treatments, genetics, phenotypes, etc. We therefore created the Mondo Disease Ontology to provide a logic-based structure for unifying multiple disease resources.

*Implications of all the available evidence:* Mondo can be leveraged by researchers and clinicians for disease annotations and data integration to aid in clinical diagnosis, treatment and advancement of human health care. Mondo is a freely available, open terminology that contains over 20,000 disease classes. Mondo is iteratively developed with contributions from the intended community and is under continuous revision, with future plans to further revise the top-level classes. Recently, efforts to classify rare diseases have centered on retrieving terms from various sources to provide a unified resource. Mondo can be explored using any of a variety of ontology browsers such as the Ontology Lookup Service (OLS) (ebi.ac.uk/ols/ontologies/mondo), and the ontology files and current releases are available on GitHub (github.com/monarch-initiative/mondo).

## Introduction

In the past decade, there have been major advances in computational approaches to disease diagnosis and care management. However, the reference data on which these tools depend are not only heterogeneous and disaggregated, but also growing and ever changing. Standard terminologies and data sources such as the Human Phenotype Ontology^1^, the Online Mendelian Inheritance in Man^2^, and Orphanet^3^ have helped standardize medical terminology for rare disease. However, reconciling the many terminologies used to name diseases and represent their inherent meaning has continued to be challenging, making knowledge and data integration difficult. It is critical to develop an unambiguous resource for disease name reconciliation such that evidence can be accurately gathered on individuals with these diseases and leveraged to inform their diagnosis, care, and treatment. This allows related resources such as gene, variant, and infectious agent resources to be interoperable and contribute to the ongoing building of medical knowledge bases.

Dozens of terminological disease resources used for research and clinical applications exist^4,5^, including for Mendelian diseases, common diseases, rare diseases, cancer, and infectious diseases, as well as others that are more comprehensive and broad^2,3,6–8^. However, scope and classification are just the beginning of the ways these resources differ: additional differences include disease naming conventions, synonym encoding, and cross references. As a result, each terminology has different strengths and weaknesses. These resources partially overlap, often significantly^9,10^ (**Figure 1**). The correspondence (mapping) among individual concepts is often accomplished through text-matching, but this can be misleading; for example, the terms ‘Muscular pseudohypertrophy-hypothyroidism syndrome’ [Orphanet:2349]^11^ and ‘B-cell immunodeficiency-limb anomaly-urogenital malformation syndrome’ [OMIM:609296]^12^ both have the exact synonym ‘Hoffman syndrome’, but they are entirely different diseases. Human-declared mappings are often represented as “cross-references”, but the relationship between the two terms can be non-exact, incorrect, out-of-date, obsolete, or otherwise not clearly defined. For example, the concept DOID:8923 ‘skin melanoma’ cross-references both OMIM:608035 ‘melanoma, cutaneous malignant, susceptibility to, 4’ and OMIM:612263 ‘melanoma, cutaneous malignant, susceptibility to, 7’^13^, which are two different types of susceptibility rather than types of melanoma. Further, some disciplines of medicine are not well covered by terminologies, for example pharmacogenetics. Therefore, the resulting integration across disease resources is often incomplete, inconsistent, and unreliable for diagnostics or research.

**Figure 1:**
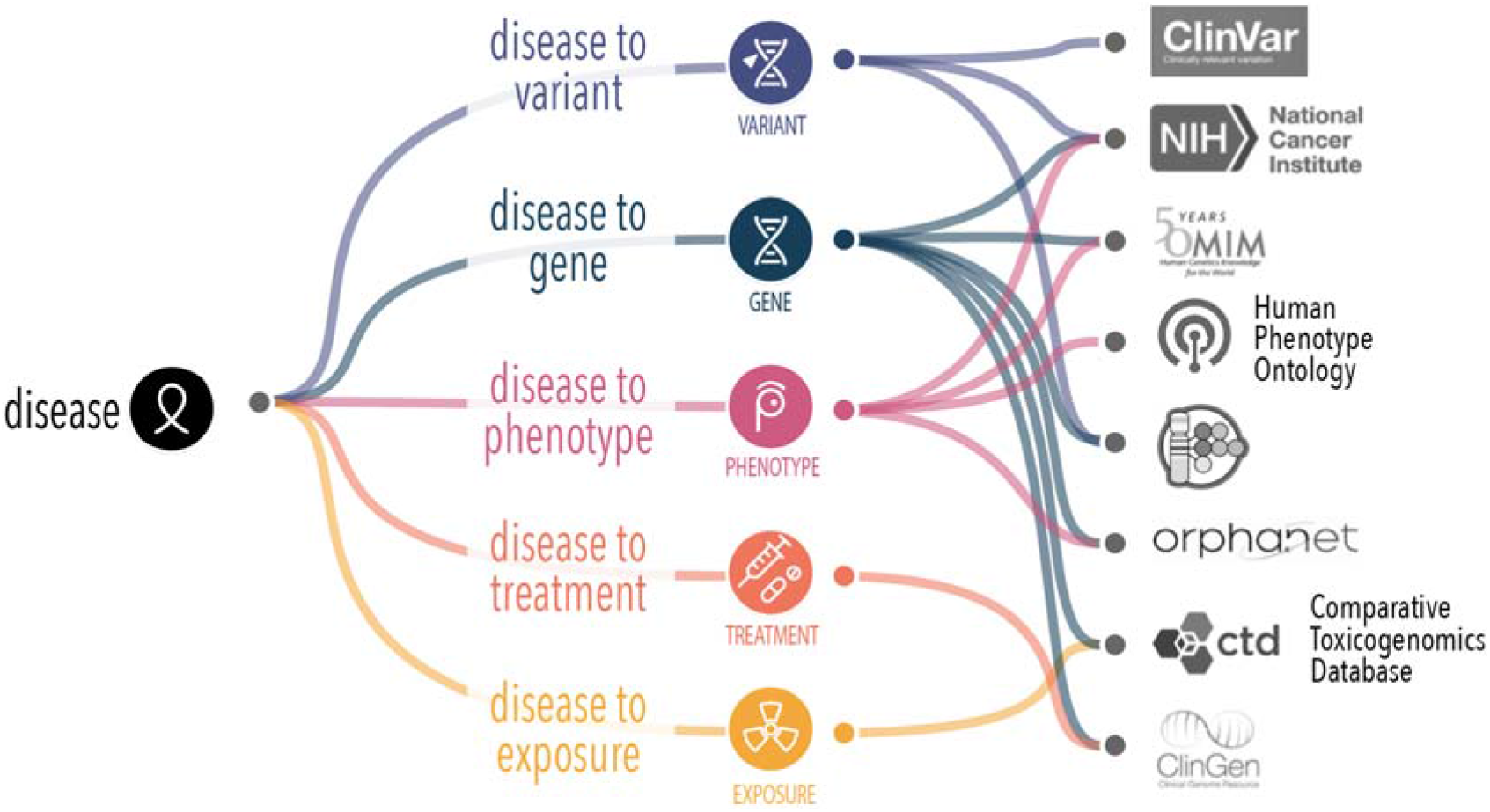
Mondo supports alignment of different disease attributes that are captured in different sources. In order to form a complete picture of knowledge about a given disease, we need an authoritative handle (stable reference) to robustly and reproducibly collate disease features.

The figure of 7,500 rare diseases^14^ is often quoted; however, in our systematic analysis across resources, we identified over 10,500 unique rare diseases^15^. Much of this heterogeneity results from a lack of consensus, both philosophically and practically, about how to classify diseases. Should diseases be classified based upon the anatomical structures they affect? Based on the doctor that first described them, such as ‘Batten disease’? Or based upon their pathogenic mechanism (e.g. infectious, deficiency, hereditary, physiological)? What if two variants in the same gene give rise to different suites of phenotypic features; are those the same disease? The ClinGen “Lumping and splitting” group (https://clinicalgenome.org/working-groups/lumping-and-splitting/) has undertaken the development of curation rules to help inform such decisions, and Orphanet has set standard procedures as well,^16^ but the community still lacks a comprehensive, multiple-parentage classification of diseases that takes into account many other features such as treatment, onset, environmental factors, to name a few. Furthermore, standard clinical enterprise terminologies such as SNOMED-CT or ICD-11 are not released with sufficient frequency to be able to keep up to date with constantly-changing disease knowledge; also, neither of these includes rarer disease codes. Combined with slow code adoption and miscoding, it continues to be very challenging to identify patients with a given disease in Electronic Health Record systems. Furthermore, numerous clinical and research systems, such as laboratory variant pipelines and repositories such as ClinVar, require up-to-date disease information. At the time the report is written or the data submitted, the disease entities should have identifiers that can be reconciled across sources and over time as knowledge changes. Fundamentally, a mechanism is needed to computationally harmonize disease classifications in order to best take advantage of our collective disease knowledge and heterogeneous data assets. This requires a modern, granular, and interoperable approach to support improved coding that can take into account the community-developed, dynamic knowledge about diseases. Here we introduce the Mondo resource, which provides a sustainable and fully-provenanced approach to integrating disease concepts from numerous sources across disease categories with the goal of better supporting precision medicine, diagnostics, and mechanistic disease research^17^.

## Mondo Disease Ontology

Terminology systems often take the form of taxonomies (simple classifications) or ontologies (conceptual domain models). The utilization of an ontology for biomedical knowledge representation enables data integration and navigation of large amounts of heterogeneous data. Additionally, an ontology encodes hierarchical and other relationships and definitions, which supports modern computational methods. Mondo includes multiple parentage, enabling concepts to be classified in multiple ways, which allows for more sophisticated querying and analytics **(Figure 2)**. For example, adult Refsum disease is a type of ‘neurometabolic disease’ and ‘phytanoyl-CoA hydroxylase deficiency’.

**Figure 2:**
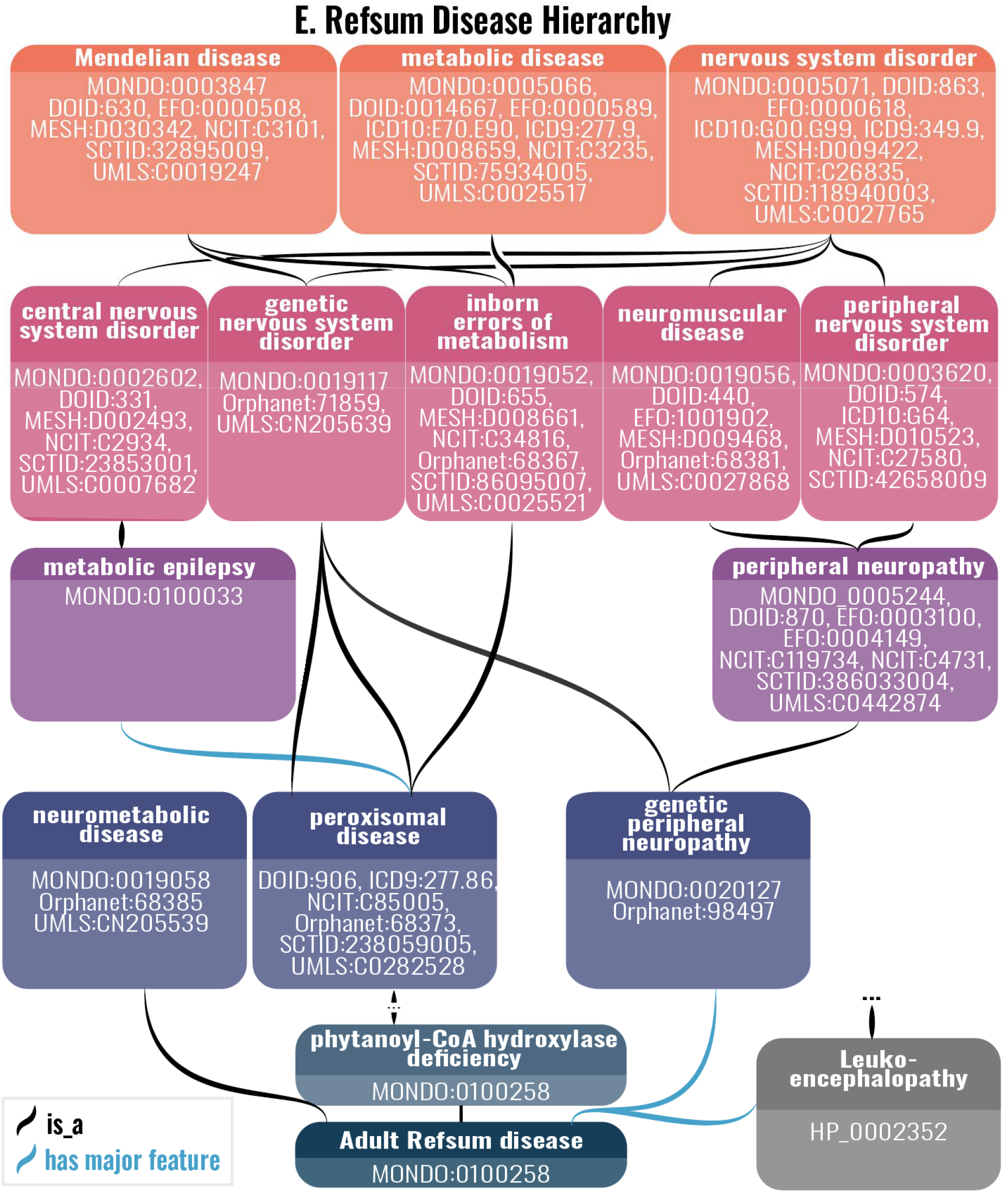
Hierarchical classification of adult Refsum disease. Mondo terms are classified in a hierarchy and can have multiple parentage, i.e., a class can have more than one parent term. Example classification of adult Refsum disease. Relationships between terms can be defined in a ‘subclass of’ relationship (is-a), or via additional relationships, such as ‘has major feature’, where a phenotype or associated disease is a feature of that disease. Each of these parent classes is similarly complex with dbxrefs spanning 10 of the 17 source terminologies in Mondo. The unique IDs and labels are shown for each term, along with the database cross-references to external ontologies (not shown for MONDO:0100258 phytanoyl-CoA hydroxylase deficiency, MONDO:0100258 Adult Refsum disease an HP:0002352 Leukoencephalopahy) Database cross-references for MONDO:0100258 Adult Refsum disease are shown in Figure 3.

Mondo currently harmonizes knowledge from 17 disease resources, collectively representing approximately 90,000 source concepts, and merges them into 22,157 distinct disease concepts (**Table 1)**. These resources were selected based on their scope, strengths, and usage (https://mondo.monarchinitiative.org/pages/sources/). Mondo covers several disease categories, including rare diseases, infectious diseases, cancers, and Mendelian diseases (**Table 2)**. Terms in Mondo have a permanent, unique identifier using the MONDO namespace, and integrate synonyms from sources that are scoped as exact, narrow, broad or related. In addition, database cross references (dbxrefs) to external sources are included, with precise semantics noting if the dbxref is equivalent or related **(Figure 3)**.

**Table 1:**
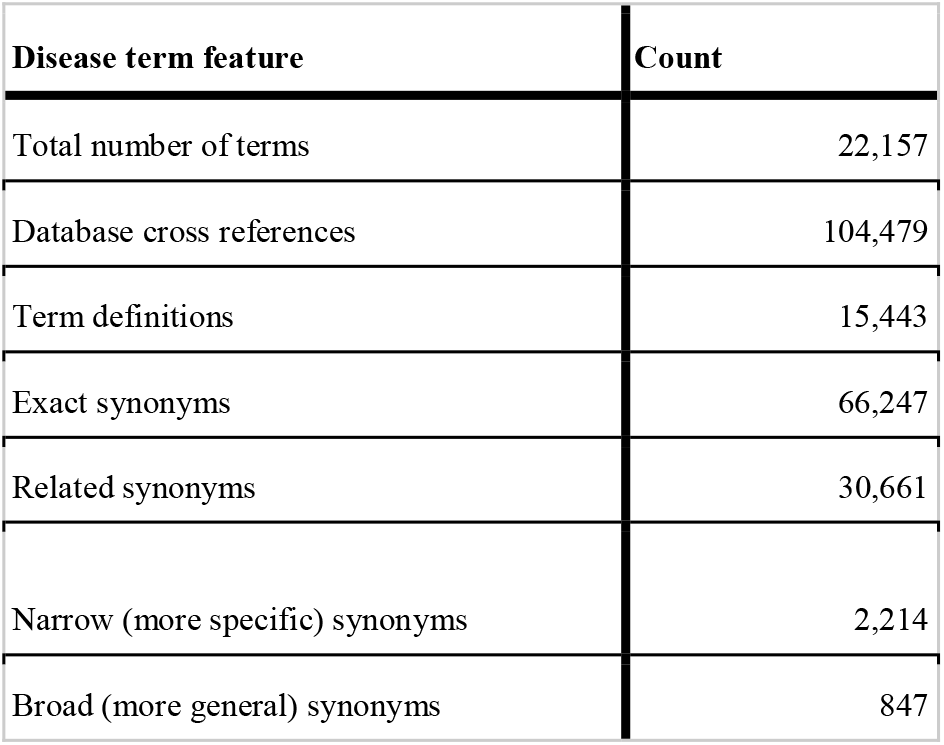
Summary statistics across all Mondo concepts. (Version at: https://github.com/monarch-initiative/mondo/releases/tag/v2022-03-01)

**Table 2:**
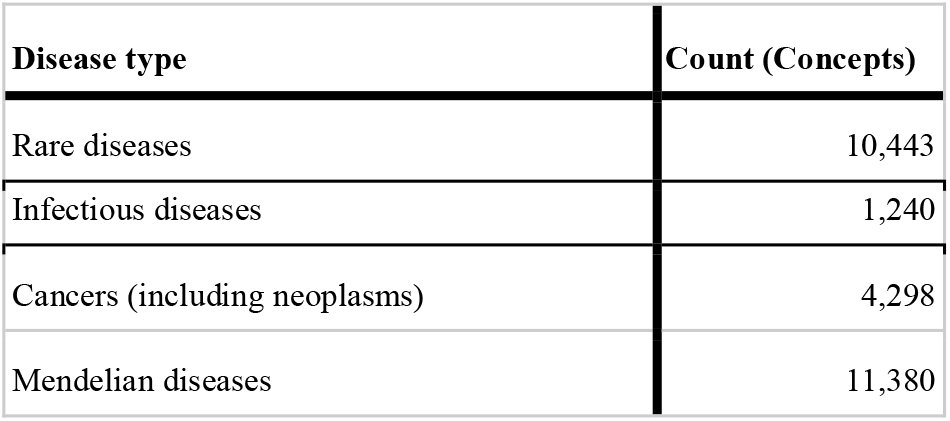
Disease concept statistics for select disease categories. Note that these groupings are overlapping. (Version at: https://github.com/monarch-initiative/mondo/releases/tag/v2022-03-01)

| Disease type | Count (Concepts) |
| --- | --- |
| Rare diseases | 10,443 |
| Infectious diseases | 1,240 |
| Cancers (including neoplasms) | 4,298 |
| Mendelian diseases | 11,380 |

**Figure 3:**
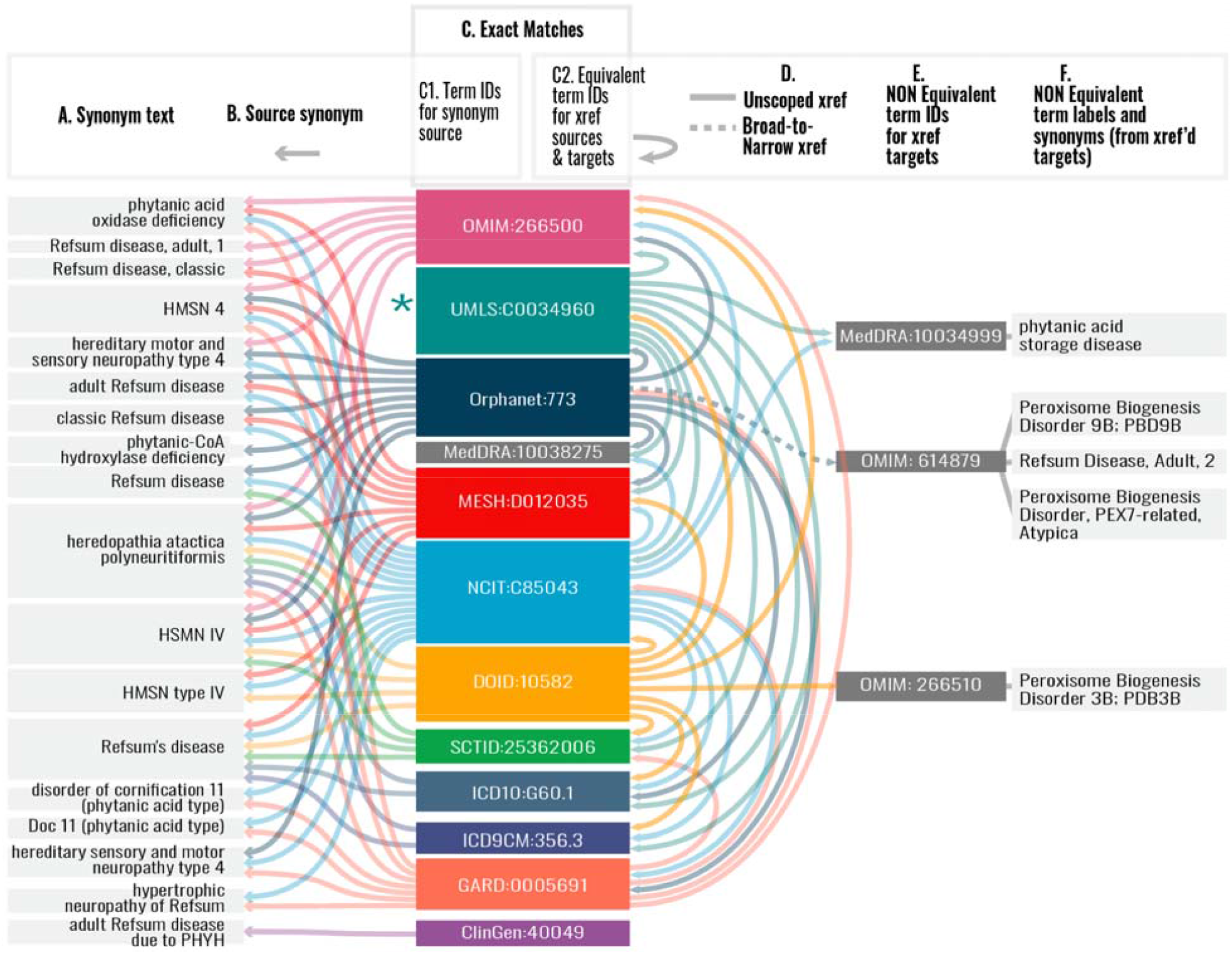
Aligning disease knowledge across sources: Mondo concept for adult Refsum disease (MONDO:0009958). A-F. A Mondo term contains synonyms scoped as exact (shown), narrow, broad and related (not shown), and database cross-references (dbxref) to the source ontologies and terminologies. A. Exact synonyms for adult Refsum disease. B. The provenance for the synonyms is captured as a database cross-reference in the Mondo ontology file. C1. Representation of the identifiers (IDs) of synonym sources, which are also database cross-references for this Mondo term. C2. Representation of the mappings between source terms and other sources. For example, UMLS:C0034960 maps to OMIM:266500. D. A solid line represents an unscoped mapping (database cross-reference, i.e. the semantics of the mapping is not defined). A dotted line represents a broad (more general) to narrow (more specific) mapping. For example, Orphanet:773 is broader than OMIM:614879. E. Represents mappings between the source term to another external term that we reviewed and determined that they are not equivalent but there is no way in the source ontologies to determine that based on the information given in the source. F. Term labels for IDs shown in E. UMLS pulls in the synonyms that are referenced by its cross-referenced neighbors (not shown). This is a subset of the mappings and does not reflect all of the mappings that exist in all of these sources.

The Human Genome Nomenclature Committee (HGNC) standardizes human gene naming, but there is no comparable global standard for reconciling the heterogeneity of disease naming systems and making them semantically interoperable. Disease names vary □— not only by language and region □— but also over time due to changing social norms and improved understanding of underlying pathogenic mechanisms; moreover, different stakeholders that speak the same language may still prefer different names for different reasons. As a consequence, it is vital to have reliable disease identifiers which durably refer to the same concept over time □— accommodating both changes and preferences. Mondo functions as a broker for disease nomenclature; the disease names and respective identifiers are a handle (i.e. stable reference), whereby synonyms, related knowledge and definitions can evolve over time with full provenance. Mondo supports multiple synonyms and synonym types as well as annotating which labels are preferred by which groups. In Mondo, synonyms are classified as exact, broad, narrow, and related ^18^. Mondo aims to accommodate all community requests and prioritizes community and medical expert recommendations for naming. More details about disease naming in Mondo is available here (https://mondo.monarchinitiative.org/pages/disease-naming/).

Articulating similarities between concepts across a set of ontologies or terminologies is challenging and often unreliable due to the prevailing use of purely automated approaches (such as text matching). Such methods lack context and can match concepts incorrectly; moreover a lack of declared rules, provenance, and versioning for these mappings makes them difficult to use for computational purposes. Mondo contains precise semantic mappings between source ontologies and terminologies, such as between OMIM, ICD10-CM, Orphanet, the National Cancer Institute Thesaurus (NCIt), and many others ^19^. A computational strategy that predicts equivalency based on a variety of features - such as labels, synonyms, cross-references (including existing semantics such as those provisioned by Orphanet), graph structure, and priors that indicate classification features specific to each source - was first applied to generate a set of mappings between concepts ^20^. The output of this computational equivalency assessment was reviewed by dedicated curation and technical teams and by the Mondo user community. Introduction of new concepts and subsequent refinements to the hierarchy and mappings are carried out as needed. Mondo will align with the new computationally friendly ICD-11, which now incorporates a pragmatic mechanism for post-coordinating terms and concepts to accommodate the granular detail of complex clinical contents.^21,22^

Mondo leverages a wealth of expert knowledge and authoritative terminologies to create a resource that is optimized for computational use in diagnostic, clinical, and research applications. Released on a monthly cycle, Mondo is iteratively developed to meet the evolving needs of a diverse, global community of contributors. There are currently more than 100 clinical and domain expert contributors from over 25 institutions that help evolve the resource, including ClinGen ^23^, OMIM ^24^, GARD ^25^, Orphanet ^26^, and others. Mondo also has a rich community of users that have implemented Mondo in a variety of settings, including incorporation into standards, such as in the Global Alliance for Genomics and Health (GA4GH) ^27^ and ISO standards such as Phenopackets ^28^, in the HL7 Terminology Authority ^29^, use in tools and data management programs such as PhenoTips^30^, as well as in databases such as ClinGen ^23^, MedGen ^31^, Gabriella Miller Kids First ^32^, Pharos ^33^, and many others. Full lists of users (https://mondo.monarchinitiative.org/pages/users/) and contributors (https://mondo.monarchinitiative.org/pages/contributors/) are available.

Mondo is more than just a source of robust and reproducible mappings between disease terminologies; Mondo also includes n-of-1, rare diseases, environmentally-influenced diseases, and complex genetic diseases that may not be documented in other sources and can be partitioned out for different uses. By integrating knowledge fully provenanced from the many existing and ever-evolving disease resources, acquired through years of work by researchers, clinicians, terminologists, and scientists from around the world, Mondo aims to make unified, comprehensive disease knowledge readily accessible to the scientific community and grow its value through logical connections across resources.

## Data Availability

All data produced are available online at https://github.com/monarch-initiative/mondo.

## Acknowledgements

Mondo is generously supported by the NIH National Human Genome Research Institute Phenomics First Resource, **NIH-NHGRI** # 1 RM1 HG010860–01, a Center of Excellence in Genomic Science; and a **NIH Office of the Director** Grant #5R24OD011883 for the Monarch Initiative. Additional support for this research/work was supported in part by the National Center for Biotechnology Information of the National Library of Medicine (NLM), National Institutes of Health. Thank you to Damien Goutte-Gattat for assistance in mining GitHub for our list of contributors.

## Data Sharing

The Mondo Disease Ontology is available at https://github.com/monarch-initiative/mondo.

## Contributor Statement

NAV and CJM directly accessed and verified the underlying data reported in the manuscript.

NAV, ST, DRU, DRM, AJM, ERR, NLH, LEC, MSK, AL, MCMT, TIO, DOS, HLR, JCS, CLT, PLW, AZ, FWN, DS, RS, IT, AHW, ALS, CGC, PNR, CJM, MAH made important intellectual contributions to manuscript revision.

NAV, DRM, AJM, ERR, LEC, MSK, AL, MCMT, TIO, DOS, HLR, JCS, CLT, PLW, AZ, GFA, JSA, LB, JPB, TIB, GAB, OJB, TJC, LCC, PCC, SLC, LCD, MD, LEF, MJF, JLG, DG, TG, MG, JAH, DSH, CTH, SK, AL, ML, ILS, BHMM, THN, DO, DPO, HP, ZMP, PR, JER, MFS, KAS, MNS, RS, RS, ALS, DS, GSS, DT, EV, DW, VW, CHX, CGC, PNR, CJM, AH, MAH contributed to development of the ontology by requesting new terms.

NAV, DRM, AJM, ERR, LEC, MSK, AL, MCMT, TIO, DOS, HLR, JCS, CLT, PLW, AZ, GFA, JSA, LB, JPB, TIB, GAB, OJB, TJC, LCC, PCC, SLC, LCD, MD, LEF, MJF, JLG, DG, TG, MG, JAH, DSH, CTH, SK, AL, ML, ILS, BHMM, THN, DO, DPO, HP, ZMP, PR, JER, MFS, KAS, MNS, RS, RS, ALS, DS, GSS, DT, EV, DW, VW, CHX, CGC, PNR, CJM, AH, MAH recommended changes to classification.

NAV, DRM, AJM, ERR, LEC, MSK, AL, MCMT, TIO, DOS, HLR, JCS, CLT, PLW, AZ, GFA, JSA, LB, JPB, TIB, GAB, OJB, TJC, LCC, PCC, SLC, LCD, MD, LEF, MJF, JLG, DG, TG, MG, JAH, DSH, CTH, SK, AL, ML, ILS, BHMM, THN, DO, DPO, HP, ZMP, PR, JER, MFS, KAS, MNS, RS, RS, ALS,

DS, GSS, DT, EV, DW, VW, CHX, CGC, PNR, CJM, AH, MAH reported bugs or other requested other changes to the ontology.

NAV, LEC, ST, PNR, CJM, AH, MAH performed data curation.

NAV, NAM, ST, JEF, HH, DRU, RS, AJM, SK, DL, JPB, EV, ALS, CJM provided technical support or quality control.

